# Abnormal Meibum Is Associated With *SREBF1* Mutation And IFAP Syndrome-2

**DOI:** 10.1101/2025.07.25.25332204

**Authors:** Igor A. Butovich, Martha Schatz, Ujwala S. Saboo, Jadwiga C. Wojtowicz, Daniel A. Johnson

**Affiliations:** University of Texas Southwestern Medical Center, Dallas, TX, USA; University of Texas Health Science Center at San Antonio, San Antonio, TX, USA

**Keywords:** Lipid metabolism, Meibomian glands, c.1579C>T gene mutation in *SREBF1*, p.Arg527Cys mutation in SREBP1, IFAP syndrome type-2, wax esters, metabolic disorders

## Abstract

The X-linked Ichthyosis Follicularis, Alopecia, and Photophobia syndrome type-2 (IFAP-2), is a condition that has been linked to a c.1579C>T mutation in the *SREBF1* gene. However, the molecular implications of the mutation in Meibomian glands (MG) remain unknown. The goals of our project were to elucidate the biochemical factors associated with IFAP-2 and develop approaches for unbiased diagnosing this condition. Meibum samples were collected from normal subjects and a patient with IFAP-2-like signs and symptoms. Genetic analysis of the IFAP-2 subject revealed a c.1579C>T (p.Arg527Cys) mutation in the SREBF-1 gene that was previously associated with IFAP-2. The meibum samples were analyzed using liquid chromatography–mass spectrometry (LC-MS), and the data were compared using multivariate statistical approaches. The LC-MS provided detailed information on the differences between the Meibomian lipid profiles of normal subjects and the IFAP-2 patient, specifically in saturated and unsaturated wax esters (SWE and UWE). Our data showed that IFAP-2 meibum was enriched with SWE which increased the SWE/UWE ratio to highly abnormal levels. The higher melting temperature of SWE compared to that of UWE correlated well with poor expressibility and abnormal thickness of IFAP-2 meibum. Thus, our study demonstrated possible links between the p.Arg527Cys mutation in SREBP1 protein, upregulation of SWE in the IFAP-2 meibum, and MG Dysfunction. It also showed that LC-MS can be used as a sensitive and informative tool to reveal minute differences in the Meibomian lipidomes of the subjects with MG Dysfunction, and identify molecular markers of the conditions.

## Introduction

Various ocular surface diseases (OSD), such as Meibomian Gland Dysfunction (MGD) and Dry Eye (DE), have a strong detrimental impact on vision and the quality of life of affected individuals (Lin et al., 2024; Valdes-Arias et al., 2024), and represent a considerable financial burden on society (Mizuno et al., 2012; Yu et al., 2011). Therefore, OSD are important targets for developing new treatments for MGD and DE. However, the multifactorial nature of MGD and DE requires targeted approaches not only for treatment, but also for reliable and objective diagnosis, which currently relies on clinical observations and testing rather than biochemical or genetic/genomic approaches. Lack of understanding of the molecular mechanisms of the onset of OSD usually prevents clinicians and biomedical scientists from developing precision therapy for patients. In many cases, OSD in general, and MGD/DE specifically, are associated with either insufficient biosynthesis and/or secretion of meibum, and its abnormal composition (Narang et al., 2023; Stapleton et al., 2025). In this manuscript, we will discuss the compositional differences between normal and IFAP-2 Meibomian gland (MG) secretions, and possible roles of the c.1579C>T gene mutation in the *SREBF1* gene on meibogenesis.

### Materials and Methods

Meibum collection procedures were approved by the Institutional Review Boards of The University of Texas Southwestern Medical Center in Dallas, TX (UTSW). The samples from an IFAP-2 female subject with severe MGD were collected at UTHSCSA as de-identified surgical discards. The collection procedures were performed in accordance with the principles of the Declaration of Helsinki. Written informed consents were obtained from the study subjects or their representatives. Standard ophthalmic approaches, such as answering OSDI questionnaire, slit lamp evaluation of the ocular surface, MG, and adnexa, and Schirmer’s tests were used to determine if the subjects had signs and symptoms of MGD/DE. Meibum samples were collected by expressing the secretion from eyelids of the subjects as described previously (Butovich, 2008). Seven normal human meibum samples were collected at UTSW, while IFAP-2 specimens were collected from a subject with severe MGD at UTHSCSA. Meibum was collected from each of the four eyelids of the IFAP-2 subject and the specimens were analyzed separately without pooling.

Meibum analysis was conducted using reverse phase liquid chromatography in combination with a high resolution atmospheric pressure chemical ionization (APCI) mass spectrometry (LC–MS) as described previously (Butovich et al., 2023). Briefly, the lipids were separated on a C18 BEH Acquity column (1mm × 100mm, 1.7μm particles) using gradient elution of the analytes with an acetonitrile/isopropanol solvent mixture. A Waters M-Class chromatograph and a high resolution time-of-flight Synapt G2-Si mass spectrometer equipped with an IonSabre-II APCI ion source (all from Waters Corp., Milford, MA) were used. The MS analyses were performed in the *m/z* (mass-to-charge) range from 100 to 2,000 arbitrary mass units with a better than ±10 mDa accuracy. The meibum lipids were detected and identified on the basis of their *m/z* values, LC retention times, and MS/MS fragmentation patterns. Where possible, the meibum lipids were authenticated using commercially available lipid standards. Each specimen was analyzed ≥5 times with a standard deviation for each sample of 3% to 7% of the corresponding means.

The data for individual meibum lipids were normalized against the total lipid content in each sample and are presented either as relative ion abundances, or as fold changes between the study samples (Butovich et al., 2023). This approach compensated for uneven physical sizes of collected meibum specimens regardless of their weights (all less than 1 mg). The data were analyzed in three different ways by conducting: 1) untargeted, unbiased, unsupervised Principal Component Analysis (PCA); 2) Orthogonal Projections to Latent Structures Discriminant Analysis (OPLS-DA) approach; and 3) targeted analyses of selected lipid species. The PCA and OPLS-DA analyses of the raw LC-MS data were performed using the Progenesis QI and EZinfo software packages, while targeted analyses of specific lipids were conducted as described before (Butovich and Wilkerson, 2022; Butovich et al., 2023) using the MassLynx/MS^E^ DataViewer software package (all from Waters Corp.). The targeted intragroup analyses of lipids were conducted using the Prism software package (version 8.0.2, from GraphPad Software, Boston, MA).

## Results and Discussion

Clinical evaluation of normal human subjects revealed no symptoms or signs of DE/MGD and no systemic diseases that would affect their vision or general lipid metabolism. The abnormal female subject was first diagnosed with OSD at the age of 1-5-yrs. The history of present illnesses included bilateral corneal opacity (Figure 1A), photophobia, debris in the tear film, and severe MGD. Subconjunctival Kenalog injections and oral Prednisolone Forte were ineffective in alleviating the conditions. At the same time, the patient had signs of cyclic alopecia in the scalp, eyebrows, and eyelashes due to keratinization, and suffered systemic skin- and hair-growth related disorders, including Netherton syndrome, trichothiodystrophy, keratosis pilaris, and atopic dermatitis. The patient was tested negative for Herpes simplex, Cytomegalovirus, and syphilis. The patient’s lipid panel was within normal limits. The results of the corneal impression cytology and lid margin cultures were also within normal limits and negative for fungal and staphylococcus infections. However, the patient had cloudy, scarred, and neovascularized corneas, and abnormally thick white tubular MG secretions in both eyes (Figure 1B) characteristic of Grade 1 MGD. Tear film breakup time measurements of were not performed.

**Figure 1.**
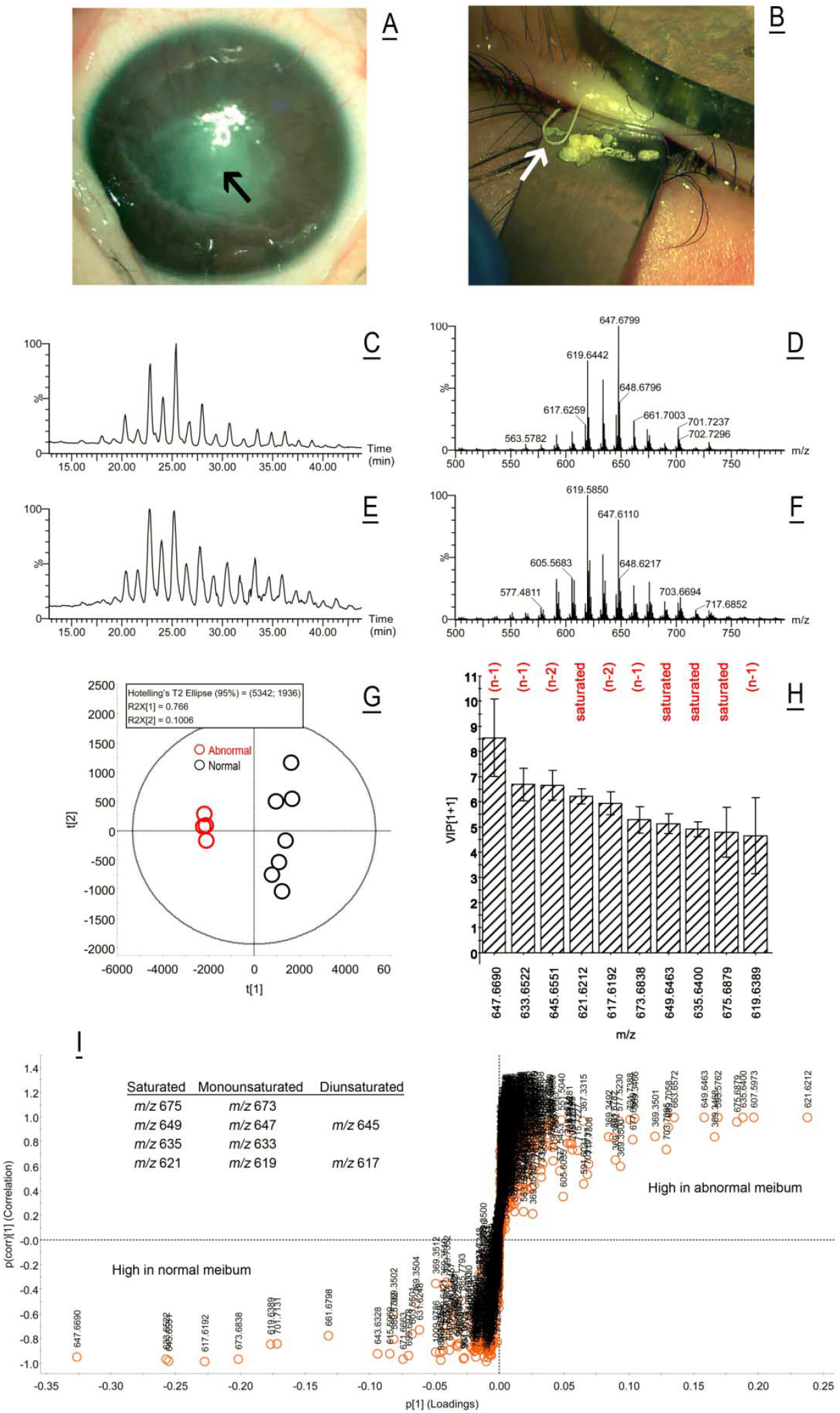
The p.Arg527Cys mutation in *SREBF1* is associated with corneal opacification, inspissated meibum, and accumulation of abnormal lipids in Meibomian gland secretions. *(<u>A</u>)* Abnormal cornea and *(<u>B</u>)* Meibomian gland secretions of the pediatric patient with a *SREBF1* mutation. The cloudy portion of the cornea and tubular Meibomian gland secretions are marked with white arrows. *(<u>C-I</u>)* Unbiased, untargeted, unsupervised lipidomic analyses of normal and IFAP-2 meibum using liquid chromatography– APCI mass spectrometry. *<u>(C</u>)* Total ion chromatogram of a representative normal human meibum sample. *(<u>D</u>*) An observation mass spectrum of the normal human meibum sample from panel A. *(<u>E</u>)* Total ion chromatogram of an abnormal IFAP-2 human meibum sample. *(<u>F</u>)* Observation mass spectrum of the abnormal human meibum sample from Panel C. *(<u>G</u>)* A PCA plot demonstrated clear separation of the abnormal samples from the normal ones. *(<u>H</u>)* A Variable Importance Plot was generated using the OPLS-DA approach and identified ten most statistically influential lipids that were responsible for differentiation of normal and abnormal study samples. *(<u>I</u>)* A S-Plot was used to identify lipids that could be considered markers of normal and abnormal meibum. Lipids in the lower right quadrant were highly expressed in normal meibum, while lipids from the upper left quadrant were overexpressed in the abnormal secretions.

Initial comparative analysis of IFAP-2 vs. normal meibum samples showed little differences between the specimens: neither their total ion chromatograms, nor observation mass spectra were visibly different (Figures 1C-1F). However, the unbiased, unsupervised, unassisted multivariate PCA demonstrated that the study samples formed two clusters (Figure 1G) – a tight cluster of individual samples from each of the four eyelids of the IFAP-2 patient and a separate cluster of seven normal subject samples – which implied the existence of substantial intergroup differences in their lipid compositions. Note that PC, which requires no prior classification of the samples and no knowledge of the number of groups, clearly separated the meibum samples into two non-overlapping clusters based on their lipidomic profiles.

To establish what the differences between the study samples might be, the data were re-analyzed using a more focused OPLS-DA approach, and a range of statistical tests was conducted. First, a Variable Importance plot (VIP) was generated, which allowed us to identify 10 most influential lipids with the greatest impact on separation of the study groups (Figure 1H). Elemental composition analysis of these lipids was performed using the EleComp software (from Waters Corp.) and suggested that they were mostly saturated and unsaturated wax esters (SWE and UWE, respectively). The identities of these analytes were verified using their LC retention times and MS/MS fragmentation patterns. The following marker WE were identified: *m/z* 617.6192– C_42_H_80_O_2_ (di-UWE); *m/z* 619.6389 – C_42_H_82_O_2_ (mono-UWE); *m/z* 621.6212 – C_42_H_84_O_2_ (SWE); *m/z* 633.6522 – C_43_H_84_O_2_ (mono-UWE); *m/z* 635.6400 – C_43_H_86_O_2_ (SWE); *m/z* 645.6551 – C_44_H_84_O_2_ (di-UWE); *m/z* 647.6690 – C_44_H_86_O_2_ (mono-UWE); *m/z* 649.6493 – C_44_H_88_O_2_ (SWE); *m/z* 673.6838 – C_46_H_90_O_2_ (mono-UWE); and *m/z* 675.6879 – C_46_H_92_O_2_ (SWE). All compounds were detected as (M + H)^+^ adducts using atmospheric pressure chemical ionization MS (APCI MS) (Butovich and Suzuki, 2020).

However, the VIP approach does not answer a question “*Which* lipids were overexpressed in *which* type of meibum?” To obtain this information, we used S-Plot analysis (Figure 1I). The lipids shown in the lower left quadrant of the graph, such as UWE, are highly expressed in the meibum of normal subjects, while those shown in the upper right quadrant, SWE, are the markers of the IFAP-2 meibum. Thus, the SWE/UWE balance was the most prominent discriminating factor between normal and IFAP-2 meibum.

Still, the S-plot did not answer another important question “What are the *fold changes* of those lipids in different types of meibum?” To address this matter, we subjected the samples to a more focused approach, targeting a few typical WE from both types of meibum (Figure 2). The results of the analyses of two representative WE – a saturated compound C_42_H_84_O_2_ and a monounsaturated C_42_H_82_O_2_ – are shown in Figures 2A1-2D2 as extracted ion chromatograms (EIC) and high resolution mass spectra. The apparent ratios of the two WE were clearly opposite in normal and IFAP-2 meibum. The apparent ratios of other SWE and UWE showed the same pattern. A comprehensive evaluation of all differences between the two types of meibum goes beyond the scope of this paper and will be reported elsewhere. For this study, we characterized the lipid specimens focusing on the gross differences between their fatty acids (FA) – essential components of WE and cholesteryl esters (CE) – using LC–MS with in-source fragmentation of lipid ions. The EIC of two major FA – palmitic acid (C_16:0_) and oleic acid (C_18:1_) – revealed the dominance of C_16:0_-based lipids in IFAP-2 meibum, and C_18:1_-based lipids in the normal one (Figures 2E1 and 2E2).

**Figure 2.**
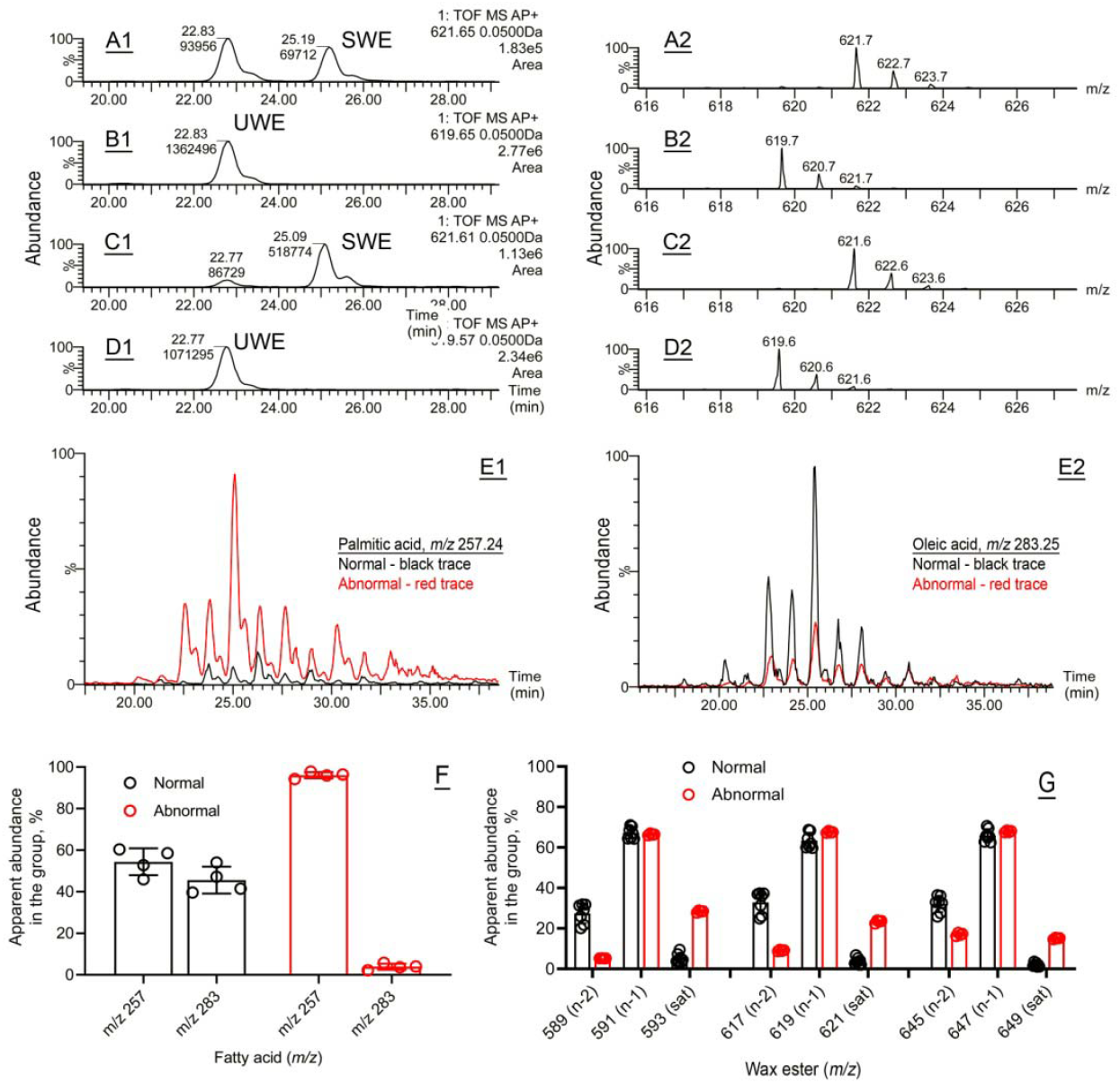
Targeted lipidomic analyses of saturated and unsaturated wax esters and fatty acids detected in normal and abnormal study samples. *(<u>A1</u>)* LC–MS detection of a saturated wax ester (SWE) C_42_H_84_O_2_ in normal meibum. The LC retention time and the peak area are shown next to the peak. *(<u>A2</u>)* The mass spectrum of the compound shown in Panel A1. *(<u>B1</u>)* Mono-unsaturated wax ester C_42_H_82_O_2_ in normal meibum. *(<u>B2</u>)* The mass spectrum of the compound shown in Panel B1. *(<u>C1</u>)* Saturated wax ester (SWE) C_42_H_84_O_2_ in abnormal meibum. *(<u>C2</u>)* The mass spectrum of the compound shown in Panel C1. *(<u>D1</u>)* Unsaturated wax ester (SWE) C_42_H_82_O_2_ in abnormal meibum. *(<u>D2</u>)* The mass spectrum of the compound shown in Panel D1. *(<u>E1</u>)* Extracted ion chromatograms of C_16:0_ palmitic acid fragments in Meibomian lipids of normal (black trace) and abnormal (red trace) meibum. The results were generated using in-source fragmentation of intact lipids in MS^E^ experiments. *(<u>E2</u>)* Extracted ion chromatograms of C_18:1_ oleic acid fragments in Meibomian lipids of normal (black trace) and abnormal (red trace) meibum. The total lipid contents of the samples were the same. *<u>(F)</u>* Balance between palmitic (*m/z* 257) and oleic (*m/z* 283) FA residues in normal and abnormal meibum. *<u>(G)</u>* Balance between saturated, monounsaturated, and diunsaturated wax esters in normal and abnormal meibum.

To quantitate the differences between our normal and IFAP-2 meibum samples, the EIC of three major FA and nine major WE were plotted, integrated and summarized to determine the apparent balances between the compounds (Figures 2F and 2G). Both experiments revealed the same trend in abnormal IFAP-2 meibum – a multifold increase in the fractions of saturated FA and WE and a decrease in unsaturated WE. Therefore, the results of our untargeted analyses of intact Meibomian lipids were supported by these targeted approaches.

Our final question was “What could the reasons for such dramatic changes in the Meibomian lipidome of the IFAP-2 subject be?”. The profound differences between the normal and the IFAP-2 Meibomian lipidomes, and the clinical signs associated with IFAP-2 were large enough to suggest that a genetic mutation in a gene that regulates lipid metabolism in MG may be responsible for this patient’s condition. Accordingly, the IFAP-2 patient’s DNA was collected, and genomic testing was performed at UTHSCSA. Full exome analysis of the patient’s DNA, revealed a CGC-to-TGC heterozygous C>T mutation in exon 8 of the *SREBF1* gene (c.1579C>T; p.Arg527Cys in SREBP1 protein).

Importantly, the c.1579C>T mutation was shown to result in various abnormal ocular and skin phenotypes, and particularly IFAP-2 (Cursiefen et al., 1999; Hamm et al., 1991; Wang et al., 2020). The mutation impaired the site-1 serine protease (S1P)-mediated cleavage of SREBP1 and its translocation to the nucleus (Wang et al., 2020), which, in turn, suppresses SCD-mediated biosynthesis of C_16:1_ palmitoleic acid (Stoffel et al., 2017). The initial clinical presentations for the IFAP-2 patient in 2018-2019 were in line with signs and symptoms of IFAP-2, while our current lipidomic data revealed the association of the mutation with abnormal meibogenesis in MG, specifically with a dramatic increase in the SWE/UWE ratios over the normal levels. It is well known that SWE have considerably higher melting temperatures than their unsaturated counterparts of the same carbon chain lengths (Patel et al., 2001). Thus, the multifold increase in the SWE/UWE ratios may be a likely reason of the anomalous IFAP-2 meibum properties reported here, which may have negative downstream impact on the ocular surface health of affected individuals.

Though treatments for IFAP-2 are yet to be developed due to its genetic nature and understudied molecular mechanisms, our findings on its association with abnormal lipid metabolism in MG provide novel insight into the physiological and biochemical impact of the mutation. To determine possible relationship between the c.1579C>T mutation in *SREBF1* and abnormal WE metabolism observed in our study, we analyzed publicly available data on potential interactions between *SREBF1* transcription factor and other lipogenic/meibogenic proteins, using the STRING (Szklarczyk et al., 2025) – an online protein-protein association and enrichment analysis database (https://string-db.org). Importantly, the protein-protein network associations of *SREBF1* include some of the major enzymes of lipogenesis, such as ACACA, FASN, and SCD, and some of the major regulatory proteins, such as INSIG1, SCAP, PPARG, PPARGC18, among others (Figure 3A). In particular, SCD is one of the major enzymes responsible for FA desaturation (Heinemann and Ozols, 2003; Ruiz et al., 2022) – a key process in maintaining a proper balance between saturated and unsaturated C_16_ and C_18_ FA, while ACACA and FASN are key enzymes for FA *de novo* biosynthesis. Taking the analysis one step further by re-centering the protein-protein interaction network on SCD, clear connections between *SREBF1*, SCD, and other enzymes of meibogenesis, such as FADS1, FADS2, SCD5, HSD17B12, and ELOVL6 (Butovich, 2017; Butovich et al., 2016) were revealed (Figure 3B), with a high confidence value of >0.7. Notably, FADS1, FADS2, and SCD5 are also FA desaturases that target C_16_ and C_18_ FA. Therefore, our results suggest that inhibition or inactivation of some (or all) of these enzymes may lead to a shift in the SWE/UWE balance as both classes of WE are largely based on saturated, mono-, and diunsaturated C_16_ to C_18_ FA (Butovich et al., 2012; Nicolaides et al., 1981). Our earlier reports on the high expression levels of *ACACA, FADS1, FADS2, FASN, ELOVL6, HSD17B12* and other related genes in human and mouse tarsal plates (Butovich, 2017; Butovich et al., 2016) reinforce this hypothesis, which is illustrated in Figure 3C and 3D.

**Figure 3.**
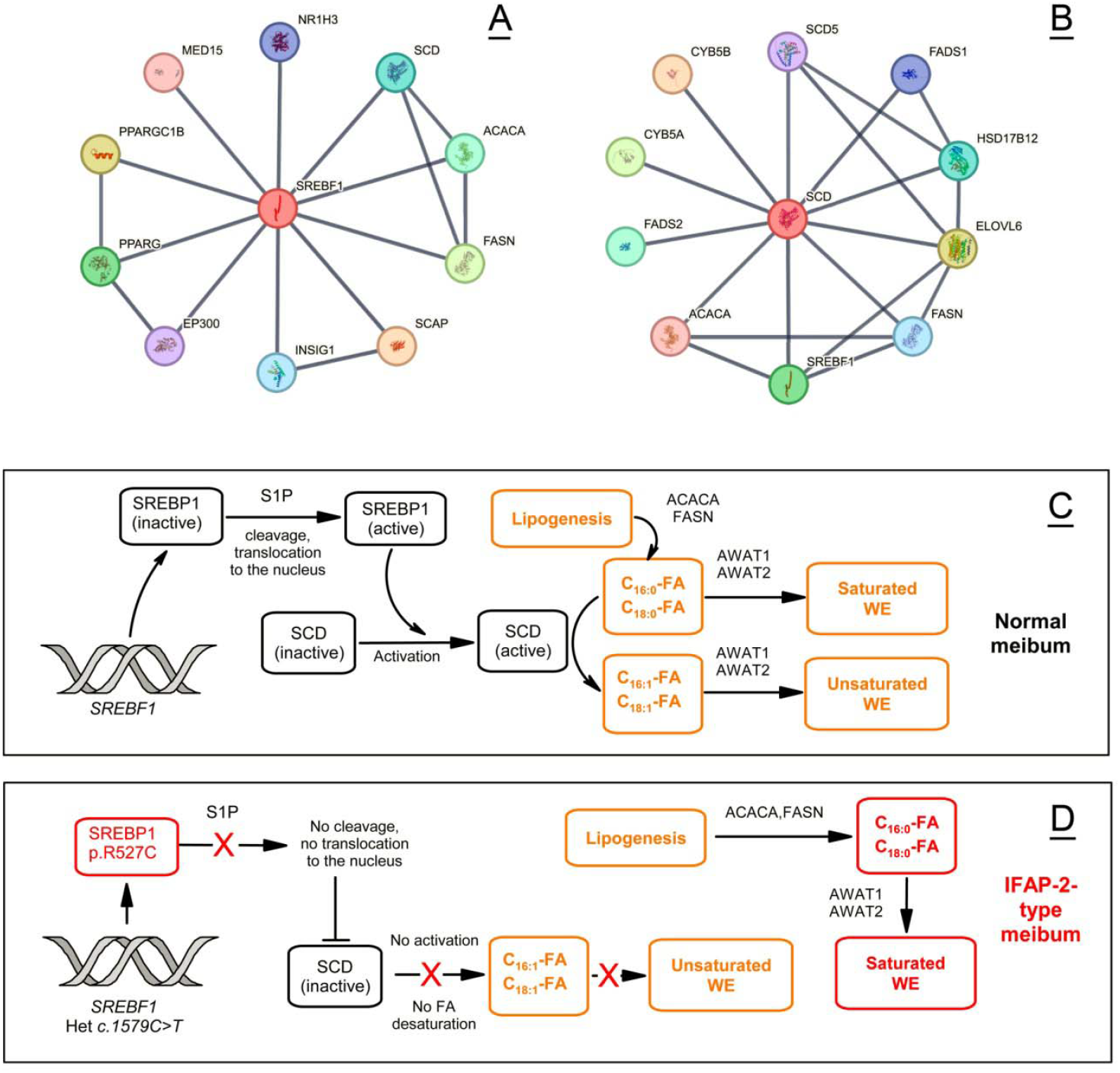
Regulation of the wax ester biosynthesis by *SREBF1* in human Meibomian glands. The STRING database was used as a source of information. The nodes represent major associated proteins that were identified in the analysis. The connecting grey lines indicate associations between the proteins. *(<u>A, B</u>)* A network of predicted and known associations for human *SREBF1*/SREBP1 *(<u>A</u>)* and SCD *(<u>B</u>)*. The following basic settings were used: Network Type – Full STRING network; Meaning of Network Edges – Confidence; Active Interaction Sources – All; Minimum Required Interaction Score – High confidence (0.700). All associations are reported with a protein-protein interaction (PPI) enrichment p-value of 1.5×10^-4^ for SREBF1, and 5.9×10^-5^ – for SCD. *(<u>C, D</u>)* Proposed mechanism of the biosynthesis of wax esters and its regulation by *SREBF1*/SREBP1 in normal Meibomian glands *(<u>C</u>)* and in the IFAP-2 subject *(<u>D</u>)*.

The main limitation of our study is that only one patient with the CGC-to-TGC heterozygous C>T mutation in the *SREBF1* gene in exon 8 (c1579C>T; p.Arg527Cys) was identified and described in this report. Due to the apparent scarcity of DNA sequencing data for individuals with DE and MGD, especially in conjunction with meibum lipid disorders, it is not known how many other reported cases of MGD are associated specifically with the shift in the SWE/UWE balance, and whether they can be traced to this, or similar, mutations in the *SREBF1* gene (Murase et al., 2021; Wang et al., 2020). We hope that our paper brings this rare condition to the attention of clinicians and researchers, leading to the recruitment of additional patients with this genetic condition and facilitating further molecular and genetic research to advance our understanding of MGD and the IFAP-2 syndrome.

In conclusion, we believe that sophisticated bioanalytical techniques such as LC–MS should be incorporated in clinical practice on a wider scale to complement clinical observations by providing valuable information on the biochemical mechanisms of pathologies and their molecular markers.

## CRediT authorship contribution statement

**I. A. Butovich:** Conceptualization, experimentation, data acquisition and analysis, writing and reviewing the manuscript, funding acquisition, supervision, project administration (at UTSW).

**M. Schatz:** Clinical testing and sample collection, data analysis, reviewing the manuscript. *U. S. Saboo: Clinical te*sting and sample collection, data analysis, reviewing the manuscript.

**J. C. Wojtowicz:** Clinical testing and sample collection, data analysis, reviewing the manuscript.

**D. A. Johnson:** Conceptualization, data acquisition, supervision, project administration, and funding acquisition (at UTHSCSA).

## Data statement

All relevant data are included in the manuscript. Additional data can be requested from the corresponding author.

